# PERSONALIZED TRANSCRANIAL ELECTRICAL STIMULATION IMPROVES SLEEP – EARLY FINDINGS

**DOI:** 10.1101/2025.10.09.25335041

**Authors:** V. Ayanampudi, A. Krishnan, R. Gurumoorthy

**Affiliations:** StimScience Inc

## Abstract

Sleep is fundamental to both physical and mental health, regulating major physiological systems (e.g., cardiovascular, immune, endocrine, metabolic) and supporting cognitive and emotional functioning here we report results of a non-invasive electrical brain stimulation approach using personalized transcranial alternating current stimulation (tACS) in improving sleep.

A total of 31 participants were recruited for null-stimulation and personalized stimulation within subjects randomized crossed design. The study was conducted with a full PSG measurement in a controlled sleep lab setting. The personalized stimulation modulated neural activity in the theta/alpha bands using a pre-sleep 15 ½ minutes stimulation. The personalization involved using each individual’s peak EEG frequencies in their theta and alpha bands.

Personalized stimulation improved sleep efficiency by 13.4%, and reduced onset latency to 6.7 minutes (a reduction of 54%), and increased the sleep duration by 26.3 minutes.

These results suggest that personalized stimulation improves sleep quality and motivates its potential for helping people suffering from insomnia.

## BACKGROUND

Sleep is fundamental to both physical and mental health, regulating major physiological systems (e.g., cardiovascular, immune, endocrine, metabolic) and supporting cognitive and emotional functioning [1]. Despite this, one in three adults sleeps less than the CDC’s recommended seven hours per night [4], and more than 30% of the general population will experience insomnia during their lifetime [25][4].

Pharmacological interventions, such as sedative hypnotics, are widely prescribed for sleep difficulties. However, these drugs have significant side effects, limited long-term efficacy, and may fail to restore normal sleep architecture [2]. Due to these safety and efficacy concerns, they are no longer recommended as a first-line treatment for insomnia [2].

These limitations have driven interest in non-pharmacological approaches to improving sleep. One such method is **non-invasive brain stimulation (NIBS)**, which uses external stimuli to modulate neural activity. Techniques include acoustic, magnetic, and electrical stimulation, each targeting outcomes such as sleep onset, quality, or duration [3].

Two primary electrical stimulation methods are **transcranial direct current stimulation (tDCS)** and **transcranial alternating current stimulation (tACS)**. Both apply low-intensity electrical currents (1–2 mA) to the scalp. While tDCS delivers a constant current, tACS alternates, enabling more flexible waveforms tailored to modulate specific neural frequency bands.

Electrical brain stimulation has a strong safety profile, with no serious adverse effects reported [13]. Large datasets—over 33,000 tDCS sessions and 1,000 participants—show only mild, transient sensations such as tingling, itching, or warmth [14][15]. tACS shares this safety record [17] and often produces even less scalp sensation than tDCS [16].

Brain oscillations occur in frequency bands tied to distinct functions [9][18][19][20][41][42][43]. Regarding sleep, EEG studies indicate that frontal-lobe oscillations are reliable biomarkers of sleep onset and maintenance [5][24], likely reflecting top-down regulation of cortico-thalamic feedback loops [6].

Experimental evidence supports the sleep-modulating potential of frontal tACS. For example, 5 Hz stimulation over the frontal cortex increases subjective sleepiness and slow-frequency EEG power [7]. Bilateral 5 Hz tACS over fronto-temporal regions can shorten sleep onset latency [30] and enhance slow-wave activity early in NREM sleep [8]. Alpha-band activity (8–13 Hz) has also been linked to drowsiness and wake–sleep transitions [31].

Past tACS sleep studies have applied uniform protocols across participants, typically targeting 0.5–16 Hz [32]. However, individuals show substantial variability in peak alpha and theta frequencies [33][34], which may limit the effectiveness of fixed protocols. This has motivated exploration of **personalized tACS**, in which stimulation is tuned to an individual’s intrinsic EEG peaks [26]. Personalized alpha or theta stimulation during wakefulness has been shown to improve entrainment, neural plasticity, and functional outcomes [35][22][23][27][12][28].

Building on this evidence, the present study tested whether personalized pre-sleep tACS could improve sleep efficiency, duration and onset. We compared two protocols: (1) a sham stimulation (30seconds ramp-up, ramp-down, followed by 15 minutes of null stimulation) and (2) a 15 ½ minute personalized waveform based on each participant’s personalized EEG peaks. Both were administered for 15 ½ minutes before bedtime using frontal electrodes. We hypothesized that personalized stimulation would outperform null-stimulation control.

## STUDY AIM

The primary aim of this study was to evaluate whether frequency-specific **personalized** transcranial alternating current stimulation (tACS) administered prior to sleep could enhance objective sleep outcomes compared to a **null-stimulation control** condition.

Specifically, the study sought to:

1. **Assess overall sleep efficiency improvement by tACS over control:** Determine whether either active stimulation protocol (Personalized) increases total sleep efficiency and reduces the tossing/turning or wake-after-sleep-onset (WASO).
2. **Compare onset latency changes with personalized vs. control protocol:** Test whether aligning stimulation frequencies to each participant’s individual EEG peaks produces superior improvements in sleep onset compared to sham control.
3. **Explore overall sleep duration changes:** Investigate whether overall sleep duration improved with the personalized stimulation.

By addressing these objectives, the study aimed to generate preliminary evidence supporting the development of individualized, non-pharmacological neuromodulation strategies to improve sleep health.

## METHOD

### Overview

Participants (n = 31; a mix of male-female, and young-old) completed a within-subject, randomized, crossover trial with two conditions: Personalized tACS, and Control (null stimulation). Single channel of stimulation using bilateral frontal electrodes (Fp1 and Fp2) delivered 15 ½ minutes of pre-sleep stimulation. Personalized stimulation frequencies were derived from individual EEG. Sham (null) stimulation was a standard 30 second ramp-up, ramp-down stimulation which was null for the remaining 15 min duration. The order of null stimulation and personalized stimulation was randomized.

Participants were outfitted with full PSG measurements including wet EEG (frontal, central, parietal, occipital electrodes), ECG, EOG and EMG (chin myography). Using a Neuroconn electrical stimulation device, we applied a single channel of stimulation using bilateral frontal electrode (Fp1 and Fp2) locations. The device used regular wet medical grade gel-based electrodes for stimulation. The stimulation waveforms of interest were programmed using their software API and uploaded into the device.

### Participants and procedures

Participants were recruited from San Francisco Bay Area. All participants were screened for neurological or medical disorders or conditions that are known to affect EEG or cognitive functioning.

A total of 31 participants were tested in a randomized, cross-over design (mean age: 56, with the age ranging between 30-70, 17 male and 14 female). Participants completed the Insomnia Severity Index questionnaire prior to their first session using an online form. Participants were primarily distributed on the first 3 ISI categories of no-insomnia, subclinical and clinical insomnia, with only 1 severe insomniac.

For the Personalized tACS condition, a preliminary session was conducted to collect EEG data from the participants to identify, on an individual basis, the power peaks within their EEG. These data were obtained during 15-minute sessions, with the participant in a relaxed, eyes-closed state, before any of the pre-sleep stimulation sessions.

The EEG signal was band pass filtered with cutoff frequencies at 0.3Hz and 45 Hz. The power spectral density (PSD) was calculated using the Welch method on the filtered data and the Fooof algorithm [18] was used to determine frequency peaks after removing the aperiodic component of the spectrum. A k-means algorithm was used to calculate power peaks within the alpha and theta bands.

For the personalized tACS condition, a stimulation waveform composed of two sinusoids was created, based on their power peaks in the alpha and theta bands. The component waveforms were started in phase and summed. These two frequency bands were targeted given their association with sleep onset, with alpha band activity (8-13Hz) linked to stage I sleep and theta band activity (4-8 Hz) linked to the transition to stage II sleep [9]. The amplitude of both sinusoids was set at 0.6mA intensity (peak-to-peak)

**Figure 1.**
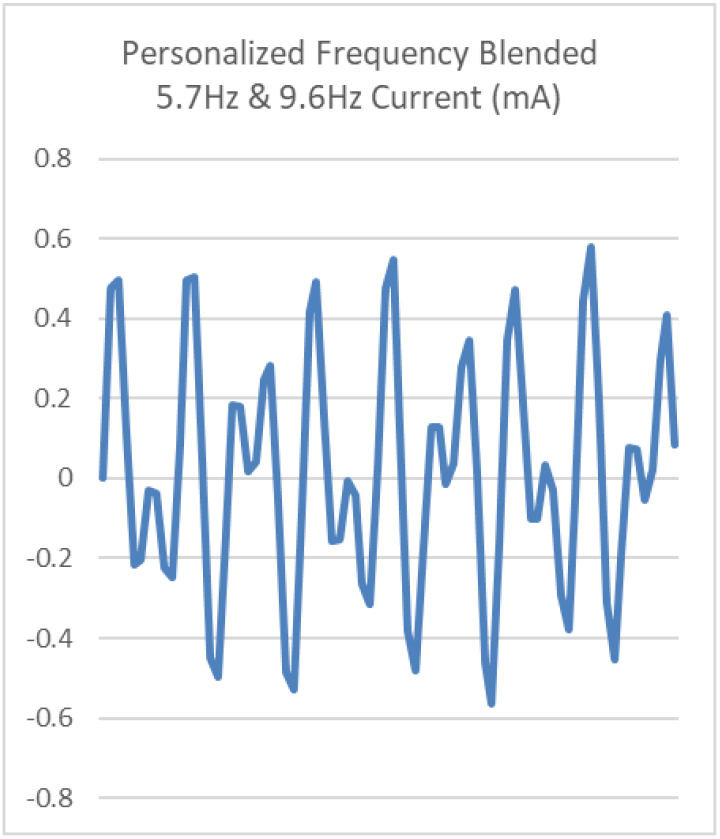
Sample personalized stimulation waveform blending 5.7Hz and 9.6Hz frequencies

### Data Collection

Continuous PSG was recorded using the NATUS PSG system, from each subject through the night with EEG (midline and bilateral coverage in the frontal, central, midline parietal and occipital, and mastoids), vertical and horizontal eye movements (EOG) were recorded using external electrodes positioned directly above and to the side of the left eye, ECG signal were also measured, along with chin myography from two electrodes on the chin. Signals were digitized and processed offline.

## DATA PREPROCESSING AND ANALYSIS

### Sleep Data Processing

The data collected was first divided into 30 second epochs. An experienced sleep scoring specialist visually inspected all the signals in each epoch and assigned a sleep stage based on predefined rules and criteria specified in the AASM manual. It was reviewed by another sleep tech separately and any differences in scoring were resolved by both scorers discussing and coming to a consensus.

### Data Analysis

Sleep tracking data was obtained using the overnight sleep tracking scores from the Natus PSG data. Sleep stage data from the device were not analyzed for this study – as the aim of the study was to explore the overview metrics of Sleep Efficiency, Onset, and Duration.

Sleep onset was defined as the interval between the end of stimulation and the first epoch scored as light, deep or REM sleep. The change in sleep onset between the two conditions was calculated as the difference in the onset of personalized stimulation compared to the null stimulation condition for each participant.

Sleep Efficiency was measured as the time slept (light sleep, deep sleep, REM) as a percentage of the time they spent in bed (including wakes in between). The change in sleep efficiency between the two conditions was calculated as the difference in the efficiency of personalized stimulation compared to the null stimulation condition as a percentage of the null stimulation measurement for each participant.

Sleep Duration was measured as the total time slept including light sleep, deep sleep, and REM sleep. The change in sleep duration between the two conditions was calculated as the difference in the sleep duration of personalized stimulation compared to the null stimulation condition.

## RESULTS

Using the PSG data, the results of the sleep measures across the personalized stimulation and sham (null-stim condition) are presented in Table 1. To get a context on the improvements seen with personalized stimulation, we reviewed other published results for sleep pill Zolpidem (Ambien) [36], and the hormonal supplement Melatonin [37],[38].

**Table 1.**
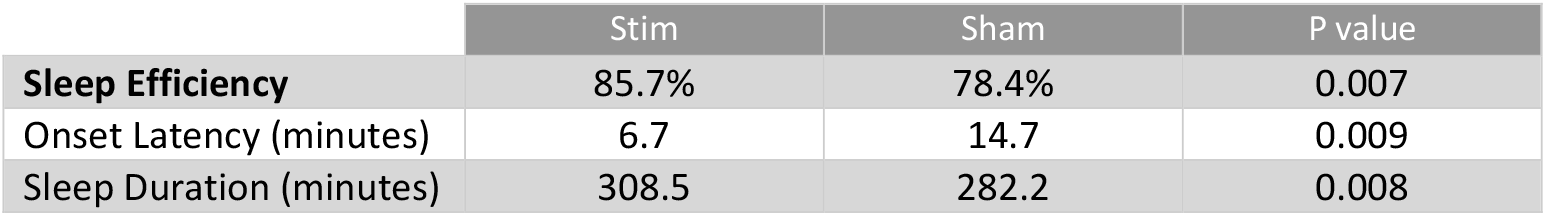
Sleep Performance Results.

### Sleep Efficiency

As seen in Table 1, personalized tACS stimulation improved sleep efficiency significantly to 85.7% from 78.4% for sham (p-value 0.007). This shows an improvement of 13.4% on average across the group. In comparison the sleeping pill Zolpidem (Ambien) reports 8.7% improvement [36], and the hormonal supplement Melatonin is the least with 3.1% improvement [37]. As we can see Personalized tACS is more efficient, almost 4x more efficient than Melatonin (Table 2).

**Table 2.** Results from this study, and reported data for Ambien, Melatonin.

|  | Sleep Efficiency<br>(% improvement) | Onset Latency<br>(mins) |
| --- | --- | --- |
| Personalized tACS | 13.4% | 6.7 |
| Ambien | 8.7% | 14.2 |
| Melatonin | 3.1% | 10.8 |

**Figure 2.**
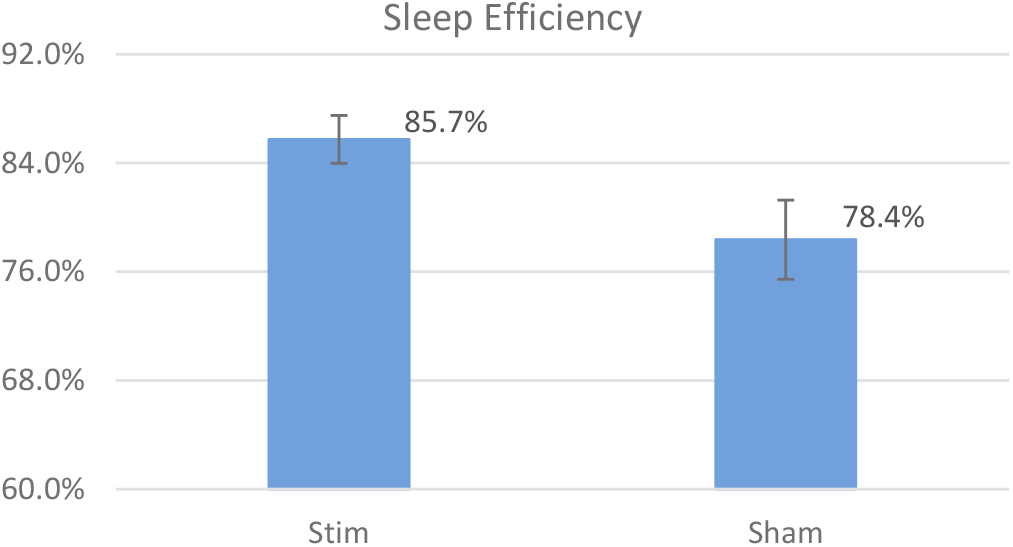
Sleep Efficiency Comparison

Looking at the sleep efficiency improvement from a perspective of the interruptions in sleep (the period spent awake in bed, also called tossing/turning) – we calculated the interruptions (WASO) as a percentage of bedtime (1 – efficiency). On the aggregate interruptions reduced from 21% to 14% (p-value 0.007) - a reduction of about a third.

### Onset Latency

Personalized tACS stimulation reduced the time taken to fall asleep to 6.7 min compared to 14.7 min for sham, a significant 8 min improvement over sham condition (p-value 0.009) (Table 1). It has cut the time taken to fall asleep by over half, across the group. In contrast, Melatonin reduces onset latency by 4 minutes [37].

**Figure 3.**
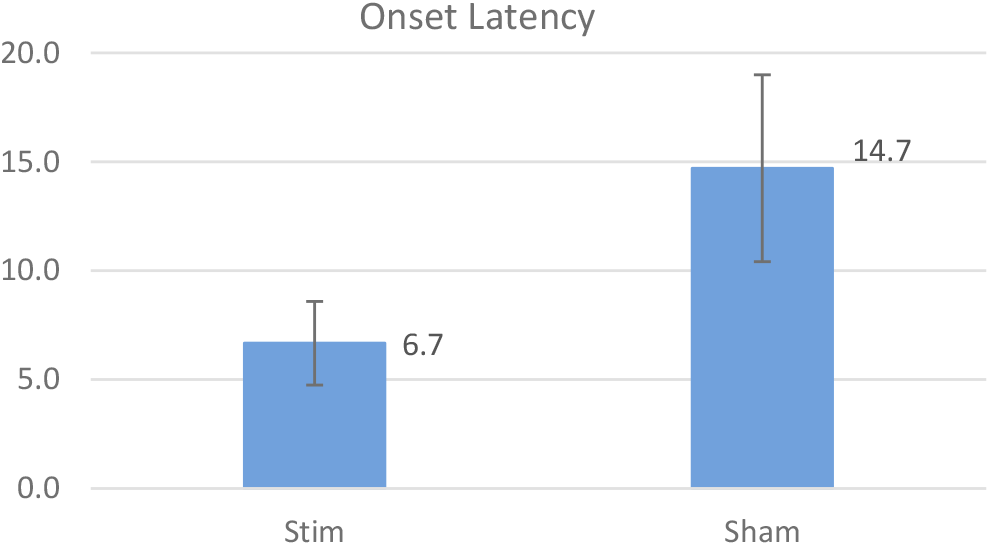
Onset Latency Comparison

### Sleep Duration

Personalized tACS stimulation added 26.3 minutes of sleep on average across the group (308.5 minutes in Stim vs. 282.2 minutes in sham, p-value 0.008). For context, Melatonin adds 12.8 minutes of sleep [37].

**Figure 4.**
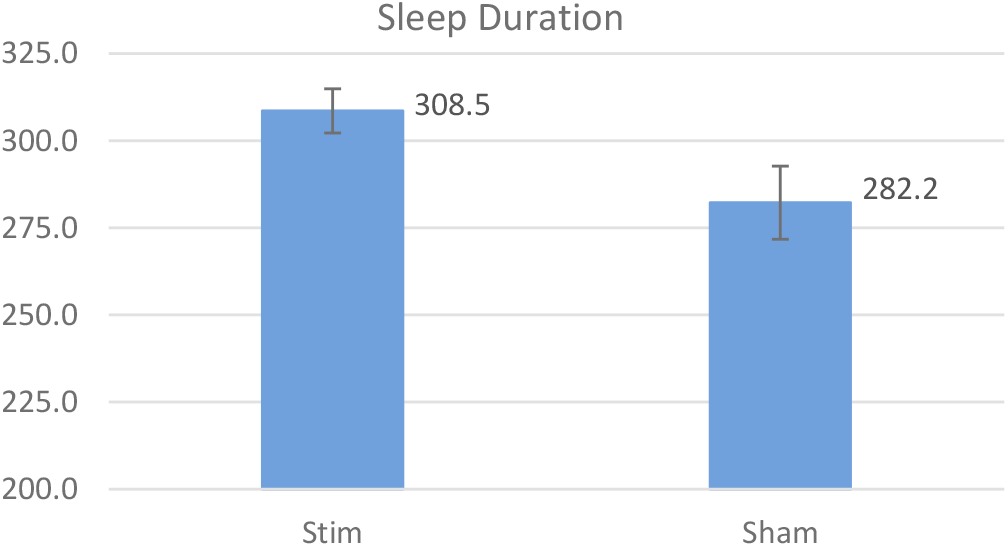
Sleep Duration Comparison

Overall Personalized tACS stimulation shows a significant improvement in sleep as measured by onset latency (8 minutes, p=0.009), sleep efficiency (13.4%, p=0.007) and sleep duration (26.3 minutes, p=0.008).

## DISCUSSION

To date, research has shown that non-invasive stimulations of varied forms can improve sleep quality [5] [6] [7] [8]. These methods include tDCS, tACS, and rTMS that modulate the electrical brain activity during sleep in an open loop i.e., fixed-stimulation manner. Studies with tACS and rTMS protocols targeting oscillatory patterns in different frequency bands (0.5Hz to 16Hz) using fixed stimulation waveforms across all subjects have shown improvements in different sleep metrics [21][39]. Additionally, invasive pharmaceutical interventions such as sleeping pills (Ambien) and Melatonin have also demonstrated varying levels of effectiveness in enhancing sleep [36][37][38].

The current study took a different approach. Specifically, the study tested whether a novel personalized tACS stimulation with multiple frequencies blended, would improve sleep. In addition, this study did stimulation before (rather than during) sleep. With participants aware of stimulation sensations, this approach has been selected based on multiple studies showing that the benefits of stimulation last well beyond the stimulation period [10][11][29].

This study has shown the efficacy of personalized tACS stimulation in improving sleep onset, duration and sleep efficiency.

A study [40], done in a user’s home setting, has explored the comparison of personalized stimulation vs fixed stimulation (using fixed 5Hz, 10Hz representation for theta and alpha bands rather than individual specific peaks). It shows that personalized tACS is indeed more efficacious in improving sleep outcomes. In addition, it showed that participants with severe insomnia benefited more from this personalized stimulation approach.

## CONCLUSIONS

Personalized electrical brain stimulation that targets specific brainwave frequencies shows promise as a way to improve sleep quality. This method could become a valuable therapeutic option in the future.

## Data Availability

All data produced in the present work are contained in the manuscript

## FUNDING

This study was internally funded by StimScience Inc. The funder had no role in study design, data collection and analysis, decision to publish, or preparation of the manuscripts.

## CONFLICTS OF INTEREST

V.Ayanampudi, A.Krishnan, and R.Gurumoorthy were employed by StimScience Inc., and have equity positions in StimScience. All authors declare no other competing interests.

## Notes

### Clinical Trial

NCT07211646

### Funding Statement

This is an internal research study funded by StimScience Inc.

### Author Declarations

Salus IRB Salus IRB Board 5 #IRB00013544 Salus IRB Study Number: 19042-05 IRB Approval This document certifies the IRB's continuing review approval and acknowledgment of the items identified under "Documents Approved" and "Documents Acknowledged" to be conducted by the named Principal Investigator. Full Board Using the criteria in 21 CFR 812.2, the FDA guidance, the IRB policies, and the information submitted, the board agreed with your contention that the device is a Non-Significant Risk (NSR) device as used in the context of this study. Please note that the FDA reserves the right to review all IRB determinations and may determine that the device is Significant Risk (SR). NOTE: Subjects must be asked for their consent using the most recently approved, stamped version(s). All IRB approved consent documents are version controlled and may not be modified in any way without prior IRB approval. Use of an unapproved document may constitute non-compliance.

### Summary of Updates

In method overview section the electrode location is Fp1-Fp2 and was originally entered as Fp3-Fp4 (which is an alternate notation for Fp1-Fp2)

